# Public awareness and support for use of wastewater for SARS-CoV-2 monitoring: A community survey in Louisville, Kentucky

**DOI:** 10.1101/2021.10.19.21264954

**Authors:** Rochelle H. Holm, J. Michael Brick, Alok R. Amraotkar, Joy L. Hart, Anish Mukherjee, Jacob Zeigler, Adrienne M. Bushau-Sprinkle, Lauren B. Anderson, Kandi L. Walker, Daymond Talley, Rachel J. Keith, Shesh N. Rai, Kenneth E. Palmer, Aruni Bhatnagar, Ted Smith

## Abstract

The majority of sewer systems in the United States and other countries, are operated by public utilities. In the absence of any regulation, public perception of monitoring wastewater for population health biomarkers is an important consideration for a public utility commission when allocating resources for this purpose. In August 2021, we conducted a survey as part of an ongoing COVID-19 community prevalence study in Louisville/Jefferson County, KY. The survey comprised of seven questions about awareness of and privacy concerns and was sent to 32,000 households randomly distributed within the county. A total of 1,220 sampled adults participated in the probability sample, and 981 were used in analysis. A total of 2,444 adults additionally responded in the convenience sample, and 1,751 were used in analysis. The samples were weighted to produce estimates representative of all adults in the county. Public awareness of tracking COVID-19 virus in the sewers was low. Opinions about how data from this activity are shared strongly supported public disclosure of monitoring results. Responses showed more support for measuring the largest areas (>30,000 to 50,000 households) typically representing population levels found in a community or regional wastewater treatment plant. Those who had a history of COVID-19 infection were more likely to support highly localized monitoring. Understanding wastewater surveillance strategies and thresholds of privacy concerns requires in-depth, comprehensive analysis of public opinion for continued success and efficacy of public health monitoring.

**Graphic for Table of Contents (TOC)/Abstract Art:** 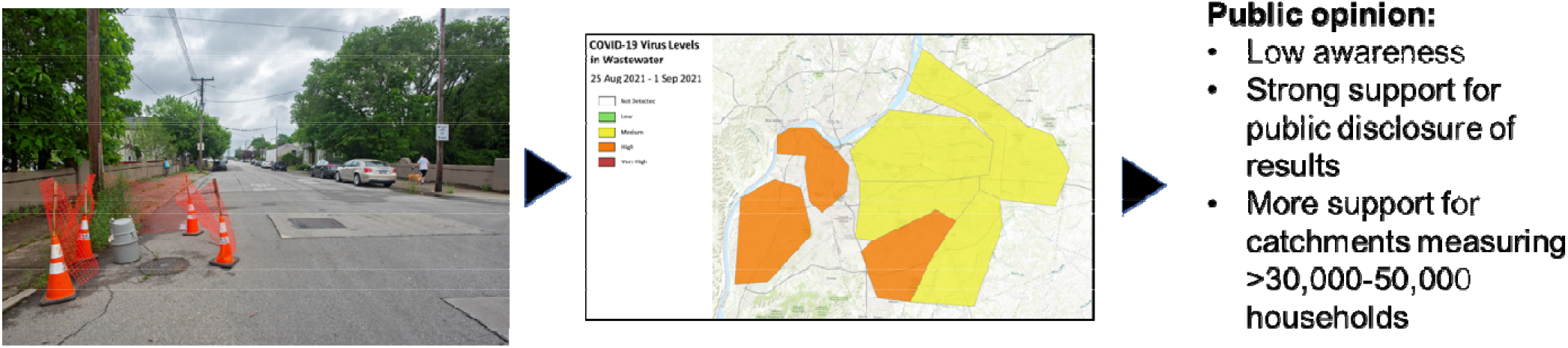

## 1. Introduction

The Coronavirus Disease 2019 (COVID-19) pandemic has brought to fore monitoring an individual’s health status related to severe acute respiratory syndrome coronavirus 2 (SARS-CoV-2) infection or vaccination. Rapid testing for the presence of the virus in the nasopharyngeal cavity and the presence of viral antigens and ant-viral antibodies in the blood have been repeatedly and widely employed. Such testing has been variably effective in preventing infections, which are primarily spread through aerosols and contaminated fluids. Additionally, individuals infected with SARS-CoV-2 also shed the virus in their stool. Therefore, rates of infection could also be estimated in the wastewater by anonymously quantifying SARS-CoV-2 genetic material in fecal matter from infected individuals who reside within an area with a piped sewer network. Abundance of the virus in wastewater has been shown to trend with infection levels measured clinically (Wu et al., 2020; Hoffmann and Alsing, 2021; Pecson et al., 2021). Globally, over 200 universities, at 2,000 sites, within 50 countries are monitoring wastewater for SARS-CoV-2 RNA (COVIDPoops19, 2021). Wastewater monitoring in the United States is being conducted by private and government laboratories, as well as academic partners; the work initiated by the United States Department of Health and Human Services alone covered wastewater SARS-CoV-2 monitoring of one-third of the US population across 42 states (Smith et al., 2021). In the United States, the Centers for Disease Control and Prevention operates a national wastewater surveillance system with SARS-CoV-2 results from which are available only to state public health officials (Centers for Disease Control and Prevention, 2021). Commercial laboratories such as Biobot Analytics have also published a national dashboard of results covering data from participating communities (https://biobot.io/data/; Biobot Analytics, Inc., 2021). Although there are ongoing legal and ethical discussions around wastewater monitoring (Gable et al., 2020; Coffman et al., 2021; Hrudey et al., 2021), the perceptions and understandings of community members whose wastewater is being monitored for SARS-CoV-2 are unknown. This information is important for future and continued application of wastewater monitoring because a majority of the sewer systems in the United States, and many other countries, are operated by public utilities. Without clearly formulated regulation, it is difficult to convince these utilities to participate in this type of sampling. In this context, public perception is an important factor that a public utility commission may need to consider when allocating resources for this purpose.

The aim of this study is to report findings on public awareness and support of SARS-CoV-2 monitoring in community wastewater from a statistically representative sample of residents in Louisville/Jefferson County, Kentucky, United States. As this line of community monitoring continues to develop, the results may inform a wider understanding of how community members monitored through an existing sewer infrastructure view public health monitoring, which may influence future approaches for disclosure and consent for wastewater surveillance and epidemiological modeling.

## 2. Methods

This study was part of a larger research project: the *Co-Immunity Project Phase II-Stratified Randomized Testing for COVID-19 Infection and Immunity in Jefferson County, KY, USA*. Study participants were 18 years and above and residents of Louisville/Jefferson County, Kentucky, United States. One group of participants was invited to enroll in the study by a postal mailing and was given an online code to consent and complete a battery of online surveys and then a few days later participated in clinical testing. This group is referred to as the probability sample. As a public service, the study was also open to all residents 18 years and older of Louisville/Jefferson County. This second group of participants, which enrolled without being invited via mail, is referred to as the convenience sample. Inclusion of the convenience sample offered a different type of population for study, and also provided an additional testing capacity for the county. The consenting and data collection procedures were identical for both probability and convenience sampling. All study participants were directed to an IRB-approved Health Insurance Portability and Accountability Act of 1996 (HIPAA) compliant secure website, where they were able to provide online signed consent, complete questionnaires, and schedule their testing appointment. Each participant provided responses to a total of 104 questions, including demographic questions, occupational information, contact and risk assessment, health history, lifestyle, COVID-19 vaccination questions, and the wastewater monitoring community survey. For this work, only demographic, COVID-19 antibody status and wastewater monitoring community survey results are reported for a single wave of this serial testing.

### 2.1. Data collection instrument

The wastewater monitoring community survey is presented in Supplement A. The survey was designed to assess the level of awareness of wastewater surveillance as a part of the COVID-19 pandemic public health response within Louisville/Jefferson County and to learn public preferences regarding how wastewater based epidemiology should be conducted. Of particular focus was the size of sewage catchment area that residents believed was appropriate for this type of health surveillance. The sewer catchment sizes, expressed as the number of households pooled in a sample, in the survey responses represented the full range of catchment areas that have been implemented in Louisville/Jefferson County (Yeager et al., 2021).

### 2.2. Serological assessment

Full methodological details for serological assessment of SARS⍰CoV⍰2 infection from this study have been recently published by Hamorsky et al. (2021). We used the antibody results from serological positivity for nucleocapsid immunoglobulin G (N-IgG) to identify participants with previous SARS-CoV-2 infection. Vaccinated respondents should not be positive for N-IgG because current COVID-19 vaccines used in the studied areas rely only on the SARS-CoV-2 viral spike protein as the immunogen.

### 2.3. Probability sampling

For the probability sample, households were contacted such that one adult within the household was randomly selected to participate. All households in Louisville/Jefferson County were stratified into 8 sectors roughly proportional to the sector size (population) based on the census block group of the address, where the area corresponded to sewer catchment areas (community sites and treatment plants). A sample of between 2,000 and 3,000 households was selected in each sector, about 32,000 total households were invited to participate in August 2021 using an address list derived from United States Postal Service delivery. In addition to the sampling strata, 4 areas (Figure 1) that were based on the demographic characteristics of the community were defined and those areas were used in the analysis (Table 1). Each selected household was mailed an invitation to participate in the study in which the sampled adult (18 years or older) was asked to complete an online informed consent, screening and survey questions and schedule an appointment for clinical testing. With the mailed invitations, each household of the probability sample population was provided with a unique personal identification registration code to be entered at the time of online registration, thereby allowing the investigators to differentiate between probability and convenience sampling populations. Each household was contacted multiple times to encourage participation. Public service announcements from the Louisville Mayor, Director of the Department of Public Health and Wellness, and mainstream media also publicized the research project.

**Table 1.**
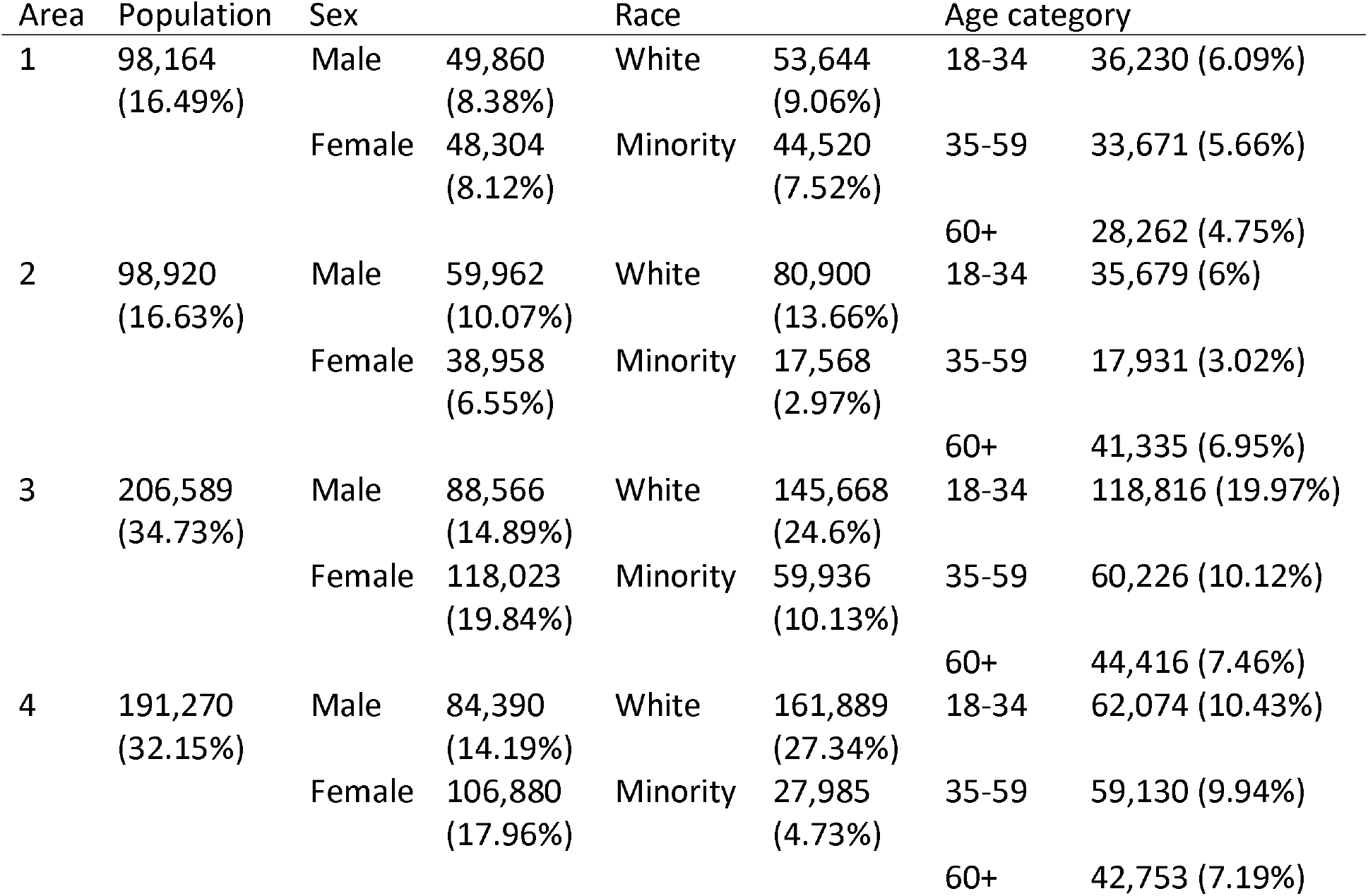
Demographic characteristics of the population surveyed.

**Figure 1.**
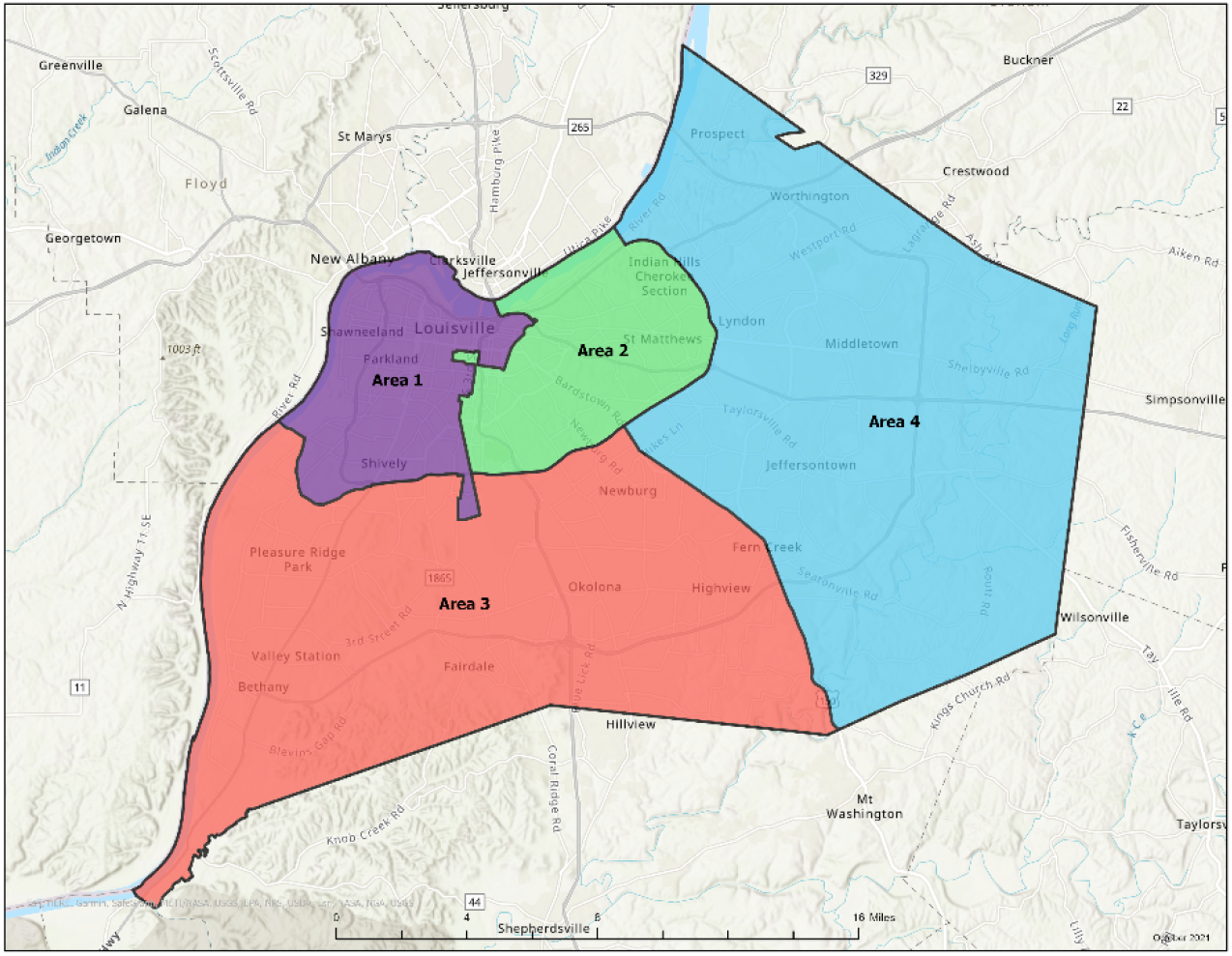
Studied area, Louisville/Jefferson County, Kentucky, United States.

### 2.4. Convenience sampling

The convenience sample was recruited using a variety of methods including social media, community outreach with organizations and influential citizens such as clergy, and public service announcements via media organizations. For example, public officials gave press conferences to publicize the efforts and local organizations made appeals to their communities. Pre-registration as well as on-site walk-up registration were both allowed.

### 2.5. Weighting the sample

The respondents were first weighted by the inverse of the probability of selection of the household and the inverse of the number of adults in the household. The final step was raking the respondents to the number of adults in the county by: sex by age, race, and geography. To produce standard errors of the estimates, 50 jackknife replicate weights were created. These replicate weights are used to estimate the standard errors of the estimates and 95 percent confidence intervals for the estimates.

### 2.6. Study participants

A total of 1,220 sampled adults participated in the probability sample, and 981 of those responded to all six multiple choice wastewater survey questions and resided in Louisville/Jefferson County and are included in this report. A total of 2,444 adults responded in the convenience sample, and 1,751 of those responded to all six multiple choice wastewater survey questions and resided in Louisville/Jefferson County are included in this report (Figure 2).

**Figure 2.**
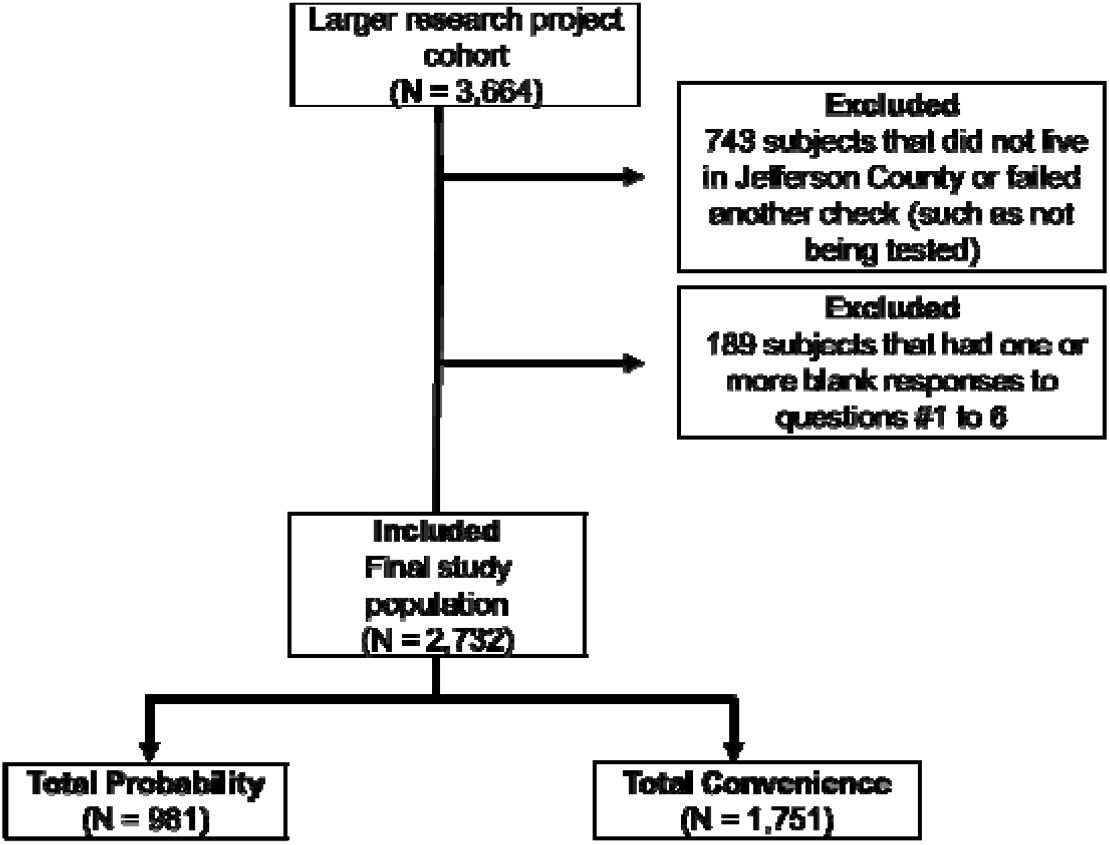
Studied population.

### 2.7. Data collection

Data were collected from August 25 to September 1, 2021.

### 2.8. Ethics

The University of Louisville Institutional Review Board approved this project as Human Subjects Research (IRB number: 20.0393 and 15.1260).

## 3. Results and discussion

### 3.1. Random probability versus convenience sample

The weighted responses from the probability and convenience samples provided estimates of the percentage of the population represented for each question. The estimates from the two samples differed substantially in several aspects (Supplement Tables B1 to B7; probability [N = 981] and convenience [N = 1,751]). Even more importantly for this analysis, the responses from the probability and convenience sample groups to the questions about wastewater monitoring varied substantially. The probability respondents were 14 to 20 percentage points less likely to indicate awareness of wastewater monitoring when compared with the convenience respondents (Supplement Table B1 to B3). Due to these differences, only the weighted random probability sample data are reported in the following quantitative analysis.

### 3.2. Wastewater monitoring awareness

When asked ‘Can the coronavirus that causes COVID-19 be detected in the city sewer system?’, 43% of respondents selected “yes”, and 49% indicated they didn’t know. More males (48%) selected “yes” than females (38%) (*p* = 0.04), and generally, an even distribution of white participants (45%) and minority participants (38%) selected “yes” (*p* = 0.18). When asked ‘Did you know that the amounts of the COVID virus in sewers reflect the general level of infection in the community?’, approximately one-third (34%) responded affirmatively. There was no difference in males (39%) that selected “yes” and females (30%) (*p* = 0.06), or for white participants (36%) and minority participants (31%) that selected “yes” (*p* = 0.21). Regarding familiarity with their wastewater utility as part of this monitoring (‘Did you know that UofL is working with Louisville Metropolitan Sewer District (MSD) to test whether measurements of coronavirus in wastewater could be used to determine the risk of COVID-19 across Louisville?’), 28% indicated that they knew MSD and UofL were conducting this monitoring.

Since the start of SARS-CoV-2 wastewater monitoring, there have been nine local news updates featuring wastewater monitoring by different media outlets in Louisville/Jefferson County (Supplement C); thus, information about the activity was shared with the public. The sewer systems in the studied area are also frequently in the news as MSD is under a Consent Decree regarding a series of sewer overflow reduction projects (MSD, 2021). Additionally, a public dashboard was initiated on May 24, 2021 to share weekly data (https://louisville.edu/envirome/thecoimmunityproject/dashboard; University of Louisville, 2021), though public engagement has been limited. And, although the national level COVIDPoops19 (2021) dashboard is available to the public, its primary audience is networking wastewater monitoring researchers.

### 3.3. Wastewater monitoring support and data sharing

The majority of respondents (85%) were supportive of wastewater sampling for public health monitoring. There was no difference in male (87%) and female (84%) participants that responded affirmatively (*p* = 0.26), while minority participants (91%) were more likely to be supportive than white participants (84%) (*p* = 0.03) (Figure 3). These results also underscore the disproportionate impact COVID-19 has had on minorities (Shiels et al., 2021) which may be driving these differences in support for public health monitoring. Importantly, in our study while some minority participants were neutral (9%), few were opposed (0.6%). Opinions about how data should be shared strongly supported (97%) public disclosure of the monitoring results. The views of male (98%) and female (97%) participants were similar (*p* = 0.46), and, although the sample size was smaller and the result was not significantly different, minority participants were unified in terms of publicly sharing such data (99%) and had ratings higher than white participants (97%) (*p* = 0.06).

**Figure 3.**
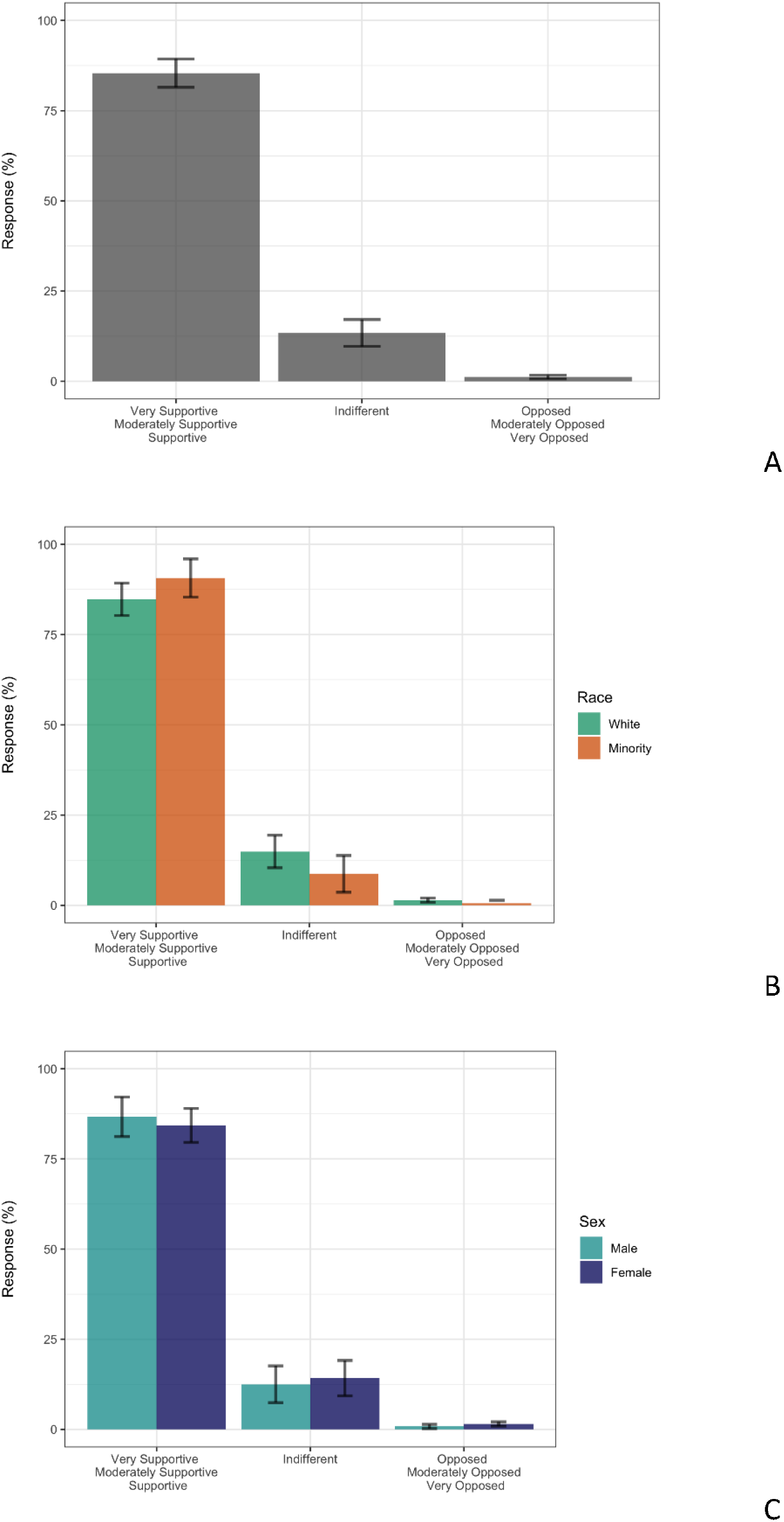
Weighted level of support for monitoring sewage to better understand COVID community infection levels instead of only testing people for probability samples (N = 981), Louisville/Jefferson County. Total survey response (A), by race (B) and by sex (C). Bars are 95% confidence intervals.

### 3.4. Size of catchment area residents believed was appropriate

The responses to the question about the smallest number of households respondents support being measured (ranging from >50,000 households to opposing any sized area) indicated considerable support (78%) for very large pooled sampling typically found in a community wastewater treatment plant (more than >50,0000 households). The next largest group (11%) indicated support for >30,000 households. The response rate of preference for wastewater monitoring at population levels >50,000 households was not different among the four areas (Rao-Scott Chi-Square Test *p* = 0.077), while the response rate of preference for community wastewater monitoring at the smallest number of households (>5,000 households) was generally lower and different (between the areas Rao-Scott Chi-Square Test *p* = 0.0008) (Figure 4). Area 1 encompasses western Louisville/Jefferson County and had the highest percentage of respondents that endorsed >50,000 households sized sampling areas. Conversely, area 4 had more variance across response options indicating a wider range of views. Area 3 has the largest portion of minority respondents in the overall study and a trend towards support for smaller, more targeted, sampling areas was observed. Opinion varies by location in the study area suggesting that there is no generic opinion for the city.

**Figure 4.**
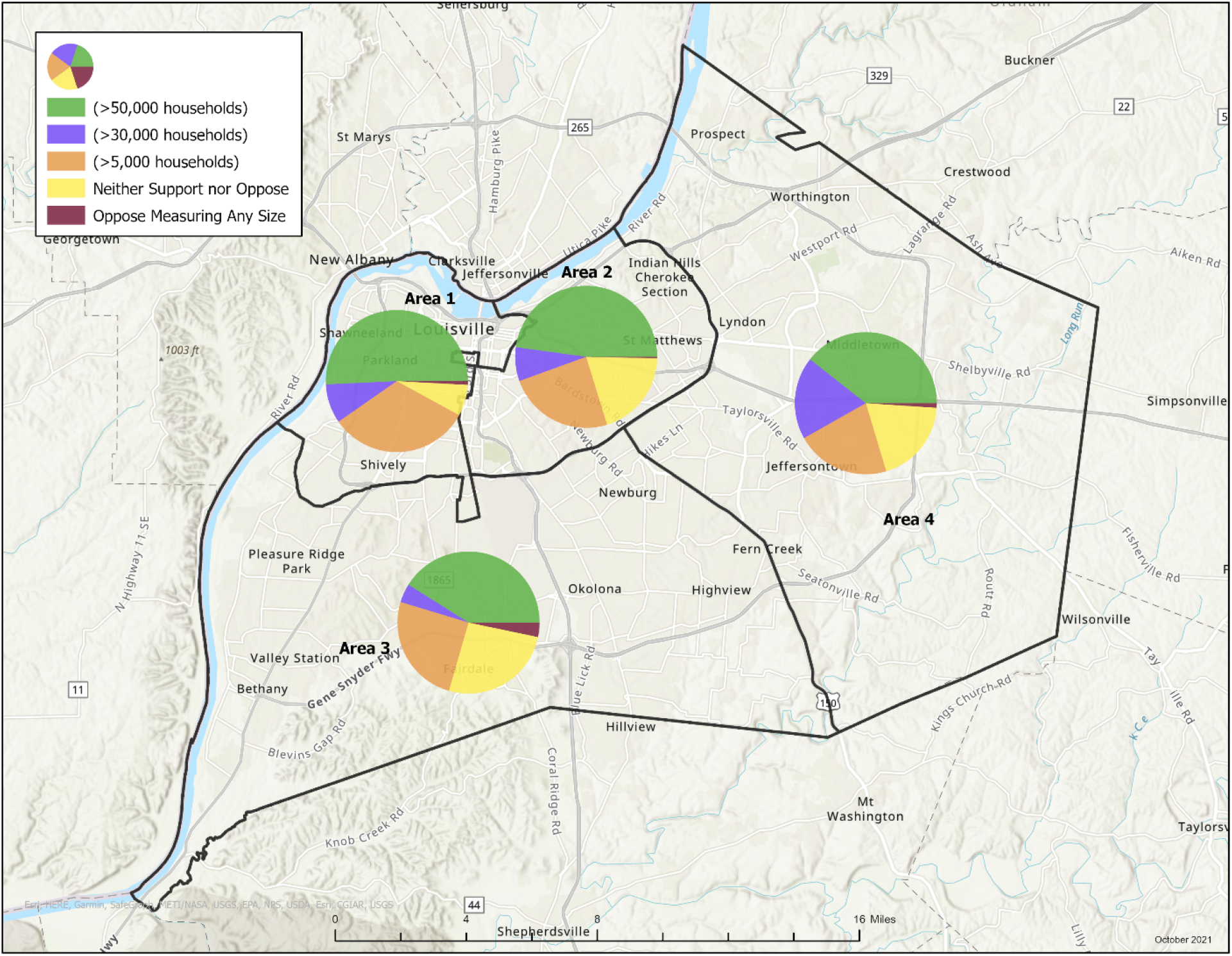
Weighted support of catchment size monitoring by geographic area for probability samples (N = 981), Louisville/Jefferson County.

An interesting finding is that support for how localized the monitoring should be varied by history of COVID-19 infection (N-IgG). Of those who had a previous infection, 42% supported the lowest threshold of 5,000 households, whereas only 22% of those who did not have a previous infection supported the smallest threshold (Figure 5). The geographic estimates of prior COVID infection were also consistent with this finding (Table 1). For example, areas 1 and 2 have previous infection rate estimates almost half of those in areas 3 and 4 (10% versus almost 20%). In the low infection areas 1 and 2, support for the highest threshold (>50,000 households) is 49%, whereas in areas 3 and 4, support for the highest threshold is 40% - almost 9 percentage points lower. How our findings relate to public awareness and support for use of wastewater monitoring outside of pandemic emergency response but related an individual’s health status for pharmaceuticals, personal care products, illicit drugs, and enteroviruses needs further study. Public opinion to surveil at the population level, and thus avoid privacy concerns, may be the most important factor to maintain the collaborative support of public utilities for such unregulated activities.

**Figure 5.**
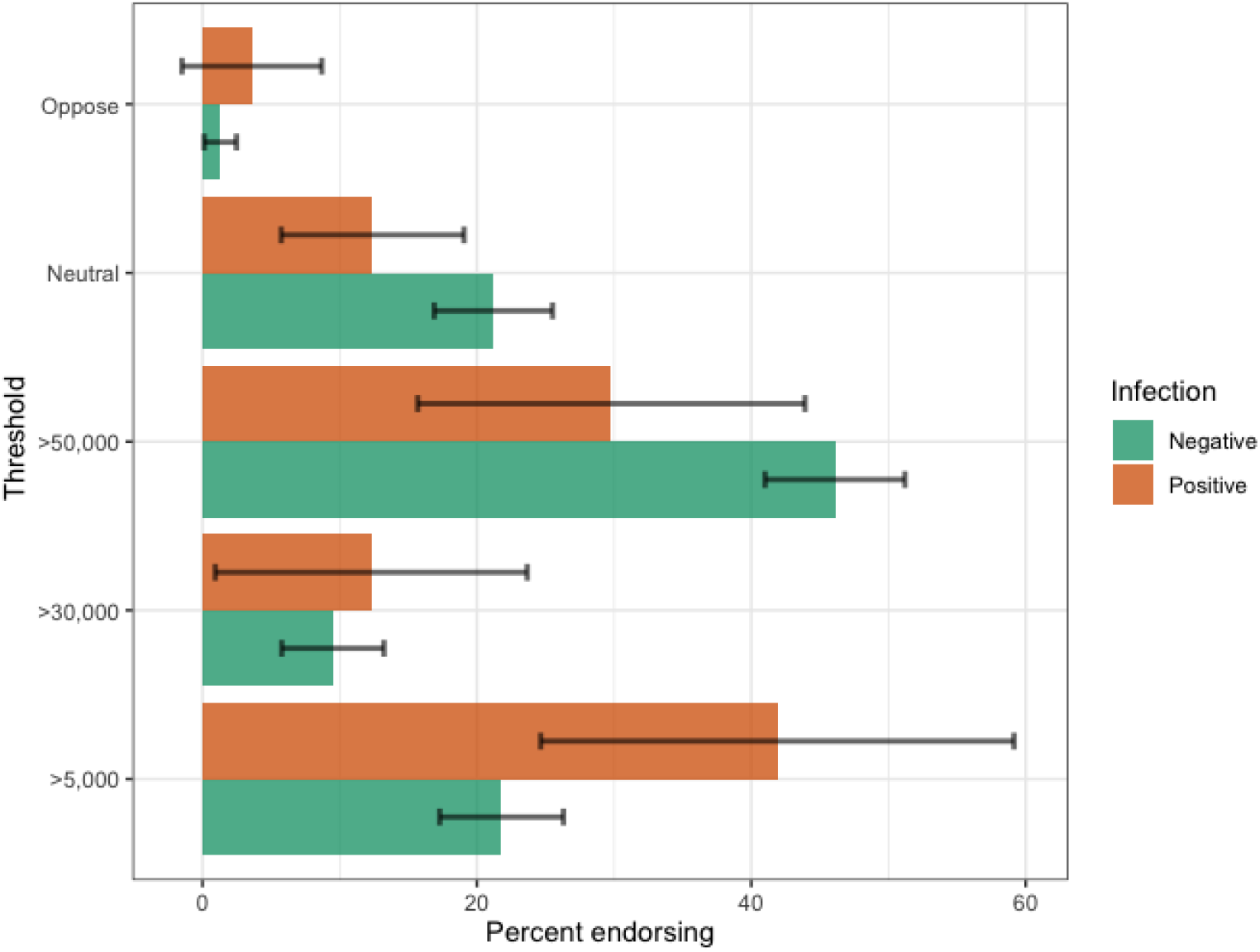
Weighted support of thresholds appropriate for wastewater monitoring by previous COVID-19 infection status (N-IgG) for probability samples (N = 981), Louisville/Jefferson County. Bars are 95% confidence intervals.

### 3.5. Qualitative feedback

We also asked for general feedback in an open-ended survey question. Of the random probability respondents ranked as being more aware and supportive of SARS-CoV-2 wastewater monitoring, some added the following comments:

> *Any way to study community spread is important, especially if people aren’t getting tested. It is a less personally invasive way of gathering that data*
>
> *-female*
>
> *If you’re monitoring for COVID, then other diseases should be monitored also…*.
>
> *-female*
>
> *The more measuring sites the better. I have no qualm with people gaining more knowledge about the health of the city. I view it no different then {than} monitoring air quality or school test scores. Feel free to do it although it sounds like gross work. Oh and thank you*.
>
> *-male*

A random probability respondent ranked as being less aware and less supportive of SARS-CoV-2 wastewater monitoring, added the following comment:

> *If such monitoring of sewers was really effective, then why have I never heard of such a thing before? There is way too much ‘false science’ combined with ‘false logic’ going around worldwide in these so called modern times. As an open minded student of science, I believe that a logic based skepticism is essential to avoid wasted time on useless pursuits*.
>
> *-male*

Only convenience sample respondent commented about sample size stating:

> *Concerned that measuring and reporting smaller areas could lead to biases based on racial, SES {*s*ocioeconomic status} or other factors. On the other hand, it could also help to get services to address health care disparities in particular areas. I’d want to really think this through if I were making a decision on this*.
>
> *-female*
>
> *As long as MSD is notifying the community that they are testing and can’t pin point a specific house I have no problem with such testing*
>
> *-male*
>
> *Workers who come into Jefferson County can bring covid and it show up in our sewers. Therefore, that area might show higher covid rates but it may not be from the people who live in that area. Being a very mobile society can skew an area or neighborhood’s results*.
>
> *-female*
>
> *I caution sponsors to avoid any focus on presumed areas of economic or social status. All results must be presented as referenced to the full community, unless specifically excluded in the project plan design*.
>
> *-female*

## 4. Limitations

Although our findings shed light on an understudied topic, the results have limitations. The large research cohort population (N = 3,664) was almost 90% vaccinated for COVID-19 in August, much higher than the, than the nearly 75 % adult residents who have received at minimum the first dose until October. Further, although a random probability and convenience sample were both used, a participant self-selection bias towards interest in research and public health is always possible.

## 5. Conclusion

Wastewater monitoring has largely been accepted as part of COVID-19 pandemic emergency response and determined to be a public health surveillance method in accordance with US Department of Health and Human Services, Title 45 Code of Federal Regulations 46, Protection of Human Subjects (US Department of Health and Human Services, 2009). Despite the likelihood our participants tended to be pro-public health, awareness overall regarding wastewater surveillance was low. Our results also underscore that, in Louisville/Jefferson County, KY, the public supports wastewater monitoring and expects to see the results of such research. We found differences in race and place across the study community which has implications for how communications about these initiatives could be improved and merits further study in other communities. Our study results suggest that to maintain public support for this type of sampling public utilities and public health professionals should consider a threshold of privacy concerns set around >30,000-50,000 households. That respondents who had a history of COVID-19 infection supported more localized monitoring suggests a possible psychographic factor which should be further explored that may account for difference in acceptance of public health activities. Despite sewers having been extensively used for public health monitoring through public utility commission participation during the COVID-19 pandemic, the use of wastewater monitoring for SARS-CoV-2 and current views of individual versus community rights, as well as privacy and informed consent, in a pandemic guarantee the issue of public awareness and support of wastewater monitoring will see increasing interest.

## Data Availability

All data produced in the present study are available upon reasonable request to the authors.

## Abbreviations

(SARS-CoV-2): severe acute respiratory syndrome coronavirus 2
(COVID-19): Coronavirus Disease 2019

## Supplement A

### Instrument: Wastewater Monitoring Community Survey

wmcs1: Can the COVID virus be detected in the city sewer system?

> 1 Yes
>
> 0 No
>
> 2 I Don’t Know

wmcs2: Did you know that the amounts of the COVID virus in sewers reflect the general level of community infection?

> 1 Yes
>
> 0 No

wmcs3: Did you know that UofL works with Louisville Metropolitan Sewer District (MSD) to study whether this kind of measurement can determine health risk across Louisville?

> 1 Yes
>
> 0 No

wmcs4: On a scale of 1 to 7, how much do you support monitoring sewage to better understand COVID infection levels in our community instead of only testing people?

> 1 Very Supportive
>
> 2 Moderately Supportive
>
> 3 Supportive
>
> 4 Indifferent
>
> 5 Opposed
>
> 6 Moderately Opposed
>
> 7 Very Opposed

wmcs5: On a scale of 1 to 7, how important is it to share what’s discovered with the public?

> 1 Very Important
>
> 2 Moderately Important
>
> 3 Supportive
>
> 4 Indifferent
>
> 5 Unimportant
>
> 6 Moderately Unimportant
>
> 7 Very Unimportant

wmcs6: Measuring at different sewer locations can help identify patterns of infection for different sized areas. Please tell us which statement best describes the smallest number of households you support being measured:

> 1 Support Measuring Largest Areas
>
> (>50,000 households)
>
> 2 Support Measuring Smaller Sections
>
> (>30,000 households)
>
> 3 Support Measuring Neighborhoods
>
> (>5,000 households)
>
> 4 Neither Support nor Oppose
>
> 5 Oppose Measuring Any Size

wmcs7: Please share any other information you’d like about your views on monitoring sewers for signs of COVID.

> (open)

## Supplement B

### Data Analysis

**Table SB 1.** Percent responding to whether the coronavirus that causes COVID-19 can be detected in the city sewer system, by sample type

| WMCS1 | Probability | Convenience |
| --- | --- | --- |
| No | 8.6 | 6.4 |
| Yes | 42.6 | 56.8 |
| Don't know | 48.8 | 36.7 |
| Total | 100.0 | 100.0 |

**Table SB 2.** Percent responding to knowing that the amounts of the COVID virus in sewers reflect the general level of infection in the community, by sample type

| WMCS2 | Probability | Convenience |
| --- | --- | --- |
| No | 65.9 | 49.5 |
| Yes | 34.1 | 50.5 |
| Total | 100.0 | 100.0 |

**Table SB 3.** Percent responding to knowing that UofL is working with Louisville Metropolitan Sewer District (MSD) to test whether measurements of coronavirus in wastewater can be used to determine the risk of COVID-19 across Louisville, by sample type

| WMCS3 | Probability | Convenience |
| --- | --- | --- |
| No | 72.4 | 52.8 |
| Yes | 27.6 | 47.2 |
| Total | 100.0 | 100.0 |

**Table SB 4.** Percent responding to how much do you support monitoring sewage to better understand COVID infection levels in our community instead of only testing people, by sample type

| WMCS4 | Probability | Convenience |
| --- | --- | --- |
| 1 | 52.2 | 61.4 |
| 2 | 11.7 | 8.1 |
| 3 | 21.5 | 14.4 |
| 4 | 13.4 | 15.0 |
| 5 | 0.5 | 0.5 |
| 6 | 0.1 | 0.1 |
| 7 | 0.6 | 0.4 |
| Total | 100.0 | 100.0 |

**Table SB 5.** Percent responding to how important is it to share the results of wastewater testing with the public, by sample type

| WMCS5 | Probability | Convenience |
| --- | --- | --- |
| 1 | 77.8 | 81.3 |
| 2 | 11.2 | 6.8 |
| 3 | 8.4 | 6.6 |
| 4 | 0.9 | 4.4 |
| 5 | 0.8 | 0.1 |
| 6 | 0.0 | 0.0 |
| 7 | 0.9 | 0.8 |
| Total | 100.0 | 100.0 |

**Table SB 6.** Percent responding to the smallest number of households support being measured, by sample type

| WMCS6 | Probability | Convenience |
| --- | --- | --- |
| 1 | 43.3 | 39.9 |
| 2 | 10.2 | 10.9 |
| 3 | 25.0 | 27.9 |
| 4 | 19.8 | 20.1 |
| 5 | 1.7 | 1.2 |
| Total | 100.0 | 100.0 |

**Table SB 7.** Percent positive for natural infection antibodies (August 2021), by sample type

| Area | Probability | Convenience |
| --- | --- | --- |
| 1 | 6.8 | 11.8 |
| 2 | 10.2 | 14.6 |
| 3 | 21.9 | 13.0 |
| 4 | 18.2 | 10.3 |

**Table SB 8.** Comparison of probability (N = 981) survey results by sex

| Male |  |  |  | Female |  |  |  |  |  |  |  |
| --- | --- | --- | --- | --- | --- | --- | --- | --- | --- | --- | --- |
| WMCS1 | n | Percent | SE | n | Percent | SE |  | diff | se(diff) | Z | p-value |
| No | 28 | 6.6 | 1.5 | 56 | 10.6 | 2.9 |  | -4.0 | 3.22 | -1.23 | 0.110 |
| Yes | 212 | 48.3 | 4.5 | 274 | 38.1 | 3.7 |  | 10.2 | 5.87 | 1.74 | 0.041 |
| DK | 160 | 45.1 | 4.0 | 251 | 51.4 | 3.6 |  | -6.3 | 5.43 | -1.15 | 0.125 |
| Total | 400 | 100.0 |  | 581 | 100.0 |  |  |  |  |  |  |
| WMCS2 | n | Percent | SE | n | Percent | SE |  |  |  |  |  |
| No | 217 | 60.8 | 4.7 | 345 | 70.0 | 3.2 |  | -9.2 | 5.75 | -1.60 | 0.055 |
| Yes | 183 | 39.2 | 4.7 | 236 | 30.0 | 3.2 |  | 9.2 | 5.75 | 1.60 | 0.055 |
| Total | 400 | 100.0 |  | 581 | 100.0 |  |  |  |  |  |  |
| WMCS3 | n | Percent | SE | n | Percent | SE |  |  |  |  |  |
| No | 268 | 76.1 | 3.5 | 395 | 69.2 | 4.7 |  | 6.9 | 5.83 | 1.17 | 0.120 |
| Yes | 132 | 23.9 | 3.5 | 186 | 30.8 | 4.7 |  | -6.9 | 5.83 | -1.17 | 0.120 |
| Total | 400 | 100.0 |  | 581 | 100.0 |  |  |  |  |  |  |
| WMCS4 | n | Percent | SE | n | Percent | SE |  |  |  |  |  |
| 1 to 3 | 331 | 86.7 | 2.8 | 486 | 84.3 | 2.4 |  | 2.4 | 3.65 | 0.65 | 0.258 |
| 4 | 64 | 12.5 | 2.6 | 87 | 14.2 | 2.5 |  | -1.7 | 3.57 | -0.48 | 0.315 |
| 5 to 7 | 5 | 0.8 | 0.3 | 8 | 1.5 | 0.3 |  | -0.7 | 0.48 | -1.37 | 0.085 |
| Total | 400 | 100.0 |  | 581 | 100.0 |  |  |  |  |  |  |
| WMCS5 | n | Percent | SE | n | Percent | SE |  |  |  |  |  |
| 1 to 3 | 385 | 97.5 | 0.8 | 568 | 97.4 | 1.3 |  | 0.1 | 1.48 | 0.09 | 0.463 |
| 4 | 9 | 1.1 | 0.5 | 7 | 0.7 | 0.5 |  | 0.4 | 0.70 | 0.56 | 0.289 |
| 5 to 7 | 6 | 1.4 | 0.7 | 6 | 1.9 | 1.1 |  |  |  |  |  |
| Total | 400 | 100.0 |  | 581 | 100.0 |  |  |  |  |  |  |
| WMCS6 | n | Percent | SE | n | Percent | SE |  |  |  |  |  |
| 1 | 143 | 35.5 | 3.7 | 252 | 50.3 | 3.2 |  | -14.8 | 4.89 | -3.02 | 0.001 |
| 2 | 42 | 12.2 | 2.5 | 55 | 8.4 | 2.3 |  | 3.7 | 3.43 | 1.09 | 0.138 |
| 3 | 117 | 33.7 | 3.9 | 124 | 17.3 | 2.4 |  | 16.5 | 4.57 | 3.61 | 0.000 |
| 4 | 91 | 16.8 | 2.7 | 145 | 22.5 | 2.6 |  | -5.7 | 3.77 | -1.50 | 0.067 |
| 5 | 7 | 1.8 | 1.0 | 5 | 1.6 | 0.9 |  | 0.2 | 1.31 | 0.17 | 0.431 |
| Total | 400 | 100.0 |  | 581 | 100.0 |  |  |  |  |  |  |

**Table SB 9.**
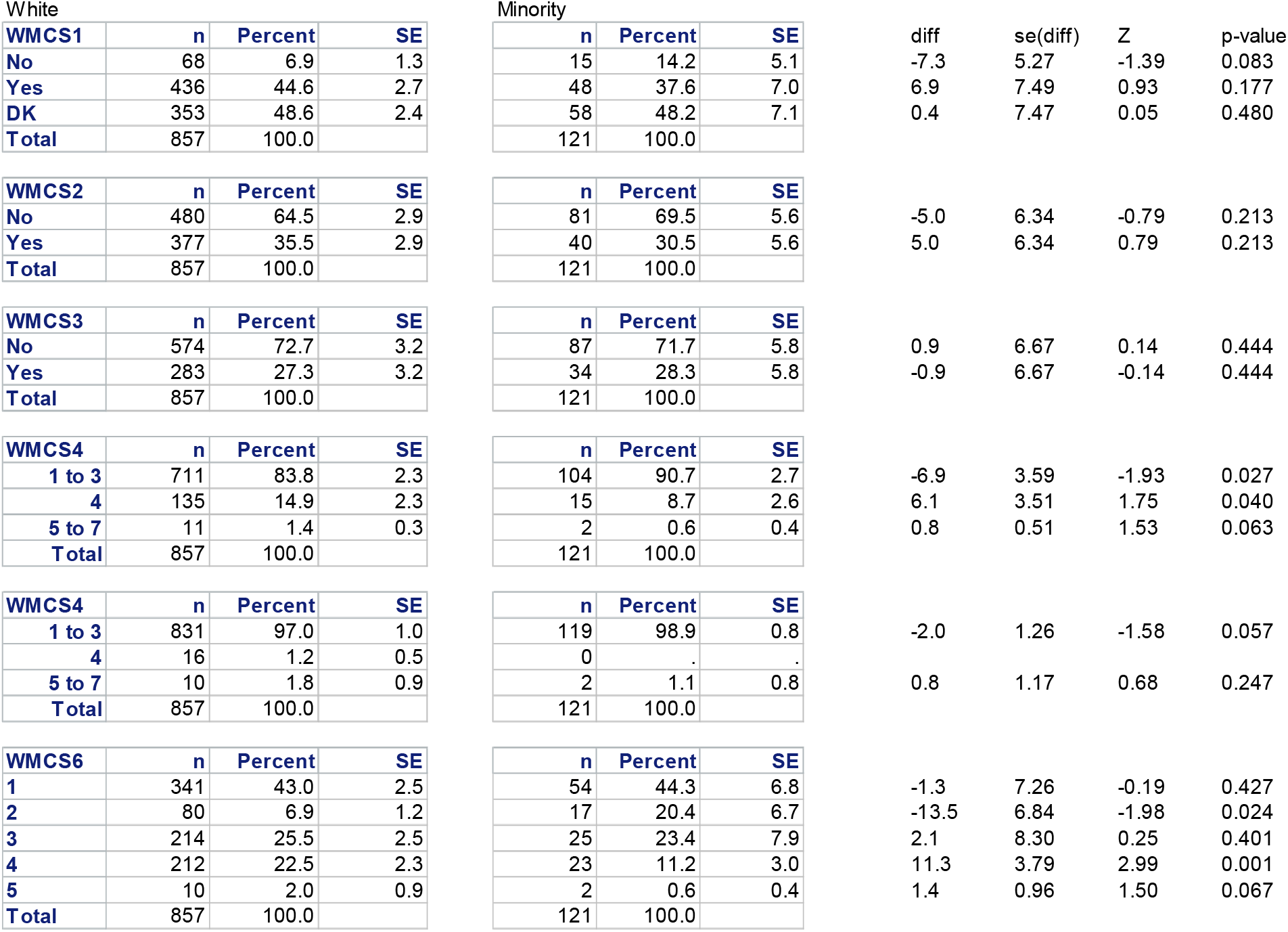
Comparison of probability (N = 981) survey results by race

**Table SB 10.** Comparison of probability (N = 981) survey results by area

| Area | WMCS1 | n | Percent | SE |  | diff-yes | se(diff) | Z | p-value |
| --- | --- | --- | --- | --- | --- | --- | --- | --- | --- |
| 1 | No | 6 | 10.4 | 6.4 | 1 vs 2 | -8.3 | 12.78 | -0.65 | 0.258 |
|  | Yes | 25 | 47.9 | 10.3 | 1 vs 3 | 10.1 | 11.09 | 0.91 | 0.181 |
|  | DK | 28 | 41.7 | 8.4 | 1 vs 4 | 9.2 | 10.98 | 0.84 | 0.200 |
|  | Total | 59 | 100.0 |  | 2 vs 3 | 18.4 | 8.68 | 2.12 | 0.017 |
| 2 | No | 16 | 6.8 | 3.8 | 2 vs 4 | 17.5 | 8.54 | 2.05 | 0.020 |
|  | Yes | 162 | 56.2 | 7.6 | 3 vs 4 | -0.9 | 5.70 | -0.15 | 0.440 |
|  | DK | 81 | 37.0 | 6.2 |  |  |  |  |  |
|  | Total | 259 | 100.0 |  |  |  |  |  |  |
| 3 | No | 21 | 8.7 | 2.1 |  |  |  |  |  |
|  | Yes | 84 | 37.8 | 4.2 |  |  |  |  |  |
|  | DK | 100 | 53.5 | 3.6 |  |  |  |  |  |
|  | Total | 205 | 100.0 |  |  |  |  |  |  |
| 4 | No | 41 | 8.8 | 2.2 |  |  |  |  |  |
|  | Yes | 215 | 38.7 | 3.9 |  |  |  |  |  |
|  | DK | 202 | 52.5 | 4.2 |  |  |  |  |  |
|  | Total | 458 | 100.0 |  |  |  |  |  |  |

| Area | WMCS2 | n | Percent | SE |  | diff-yes | se(diff) | Z | p-value |
| --- | --- | --- | --- | --- | --- | --- | --- | --- | --- |
| 1 | No | 39 | 70.1 | 7.8 | 1 vs 2 | -17.0 | 11.02 | -1.54 | 0.061 |
|  | Yes | 20 | 29.9 | 7.8 | 1 vs 3 | -1.0 | 8.86 | -0.12 | 0.453 |
|  | Total | 59 | 100.0 |  | 1 vs 4 | -3.8 | 8.77 | -0.44 | 0.331 |
| 2 | No | 115 | 53.1 | 7.8 | 2 vs 3 | 16.0 | 8.89 | 1.80 | 0.036 |
|  | Yes | 144 | 46.9 | 7.8 | 2 vs 4 | 13.2 | 8.80 | 1.50 | 0.067 |
|  | Total | 259 | 100.0 |  | 3 vs 4 | -2.8 | 5.87 | -0.47 | 0.318 |
| 3 | No | 137 | 69.1 | 4.2 |  |  |  |  |  |
|  | Yes | 68 | 30.9 | 4.2 |  |  |  |  |  |
|  | Total | 205 | 100.0 |  |  |  |  |  |  |
| 4 | No | 271 | 66.3 | 4.1 |  |  |  |  |  |
|  | Yes | 187 | 33.7 | 4.1 |  |  |  |  |  |
|  | Total | 458 | 100.0 |  |  |  |  |  |  |

| Area | WMCS3 | n | Percent | SE |  | diff-yes | se(diff) | Z | p-value |
| --- | --- | --- | --- | --- | --- | --- | --- | --- | --- |
| **1** | **No** | 41 | 62.5 | 9.9 | 1 vs 2 | 4.3 | 12.22 | 0.35 | 0.364 |
|  | **Yes** | 18 | 37.5 | 9.9 | 1 vs 3 | 13.5 | 10.83 | 1.25 | 0.106 |
|  | **Total** | 59 | 100.0 |  | 1 vs 4 | 14.4 | 10.32 | 1.40 | 0.081 |
| **2** | **No** | 152 | 66.7 | 7.1 | 2 vs 3 | 9.2 | 8.35 | 1.11 | 0.135 |
|  | **Yes** | 107 | 33.3 | 7.1 | 2 vs 4 | 10.2 | 7.68 | 1.32 | 0.093 |
|  | **Total** | 259 | 100.0 |  | 3 vs 4 | 0.9 | 5.19 | 0.18 | 0.429 |
| **3** | **No** | 144 | 76.0 | 4.3 |  |  |  |  |  |
|  | **Yes** | 61 | 24.0 | 4.3 |  |  |  |  |  |
|  | **Total** | 205 | 100.0 |  |  |  |  |  |  |
| **4** | **No** | 326 | 76.9 | 2.8 |  |  |  |  |  |
|  | **Yes** | 132 | 23.1 | 2.8 |  |  |  |  |  |
|  | **Total** | 458 | 100.0 |  |  |  |  |  |  |

| Area | WMCS4 | n | Percent | SE |  | diff-1-3 | se(diff) | Z | p-value |
| --- | --- | --- | --- | --- | --- | --- | --- | --- | --- |
| **1** | **1 to 3** | 53 | 87.2 | 6.0 | 1 vs 2 | 1.3 | 6.92 | 0.19 | 0.425 |
|  | **4** | 5 | 11.9 | 5.6 | 1 vs 3 | 3.5 | 6.83 | 0.52 | 0.302 |
|  | **5 to 7** | 1 | 0.9 | 0.6 | 1 vs 4 | 1.1 | 6.45 | 0.17 | 0.432 |
|  | **Total** | 59 | 100.0 |  | 2 vs 3 | 2.2 | 4.77 | 0.47 | 0.320 |
| **2** | **1 to 3** | 215 | 85.9 | 3.5 | 2 vs 4 | -0.2 | 4.21 | -0.05 | 0.481 |
|  | **4** | 41 | 12.3 | 3.3 | 3 vs 4 | -2.4 | 4.06 | -0.60 | 0.274 |
|  | **5 to 7** | 3 | 1.7 | 1.4 |  |  |  |  |  |
|  | **Total** | 259 | 100.0 |  |  |  |  |  |  |
| **3** | **1 to 3** | 162 | 83.7 | 3.3 |  |  |  |  |  |
|  | **4** | 39 | 15.0 | 3.4 |  |  |  |  |  |
|  | **5 to 7** | 4 | 1.3 | 0.7 |  |  |  |  |  |
|  | **Total** | 205 | 100.0 |  |  |  |  |  |  |
| **4** | **1 to 3** | 387 | 86.1 | 2.4 |  |  |  |  |  |
|  | **4** | 66 | 12.9 | 2.3 |  |  |  |  |  |
|  | **5 to 7** | 5 | 0.9 | 0.4 |  |  |  |  |  |
|  | **Total** | 458 | 100.0 |  |  |  |  |  |  |

| Area | WMCS5 | n | Percent | SE |  | diff-1-3 | se(diff) | Z | p-value |
| --- | --- | --- | --- | --- | --- | --- | --- | --- | --- |
| **1** | **1 to 3** | 59 | 100.0 | 0.0 | 1 vs 2 | 3.2 | 1.67 | 1.93 | 0.027 |
|  | **4** | 0 | . | . | 1 vs 3 | 4.7 | 1.98 | 2.38 | 0.009 |
|  | **5 to 7** | 0 | . | . | 1 vs 4 | 1.1 | 0.52 | 2.20 | 0.014 |
|  | **Total** | 59 | 100.0 |  | 2 vs 3 | 1.5 | 2.59 | 0.58 | 0.281 |
| **2** | **1 to 3** | 250 | 96.8 | 1.7 | 2 vs 4 | -2.1 | 1.75 | -1.19 | 0.117 |
|  | **4** | 4 | 1.7 | 1.4 | 3 vs 4 | -3.6 | 2.05 | -1.75 | 0.040 |
|  | **5 to 7** | 5 | 1.5 | 0.9 |  |  |  |  |  |
|  | **Total** | 259 | 100.0 |  |  |  |  |  |  |
| **3** | **1 to 3** | 193 | 95.3 | 2.0 |  |  |  |  |  |
|  | **4** | 7 | 1.3 | 0.6 |  |  |  |  |  |
|  | **5 to 7** | 5 | 3.4 | 1.9 |  |  |  |  |  |
|  | **Total** | 205 | 100.0 |  |  |  |  |  |  |
| **4** | **1 to 3** | 451 | 98.9 | 0.5 |  |  |  |  |  |
|  | **4** | 5 | 0.5 | 0.3 |  |  |  |  |  |
|  | **5 to 7** | 2 | 0.6 | 0.5 |  |  |  |  |  |
|  | **Total** | 458 | 100.0 |  |  |  |  |  |  |

| Area | WMCS6 | n | Percent | SE |  | diff-1 | se(diff) | Z | p-value |
| --- | --- | --- | --- | --- | --- | --- | --- | --- | --- |
| **1** | **1** | 28 | 50.8 | 9.8 | 1 vs 2 | 2.9 | 11.87 | 0.25 | 0.403 |
|  | **2** | 5 | 8.9 | 6.3 | 1 vs 3 | 9.7 | 10.91 | 0.89 | 0.187 |
|  | **3** | 18 | 32.3 | 9.5 | 1 vs 4 | 11.3 | 10.33 | 1.10 | 0.136 |
|  | **4** | 7 | 7.1 | 3.7 | 2 vs 3 | 6.8 | 8.32 | 0.81 | 0.208 |
|  | **5** | 1 | 0.9 | 0.6 | 2 vs 4 | 8.4 | 7.55 | 1.12 | 0.132 |
|  | **Total** | 59 | 100.0 |  | 3 vs 4 | 1.7 | 5.92 | 0.28 | 0.390 |
| **2** | **1** | 105 | 47.8 | 6.8 |  |  |  |  |  |
|  | **2** | 23 | 7.7 | 2.9 |  |  |  |  |  |
|  | **3** | 69 | 24.2 | 4.6 |  |  |  |  |  |
|  | **4** | 61 | 19.9 | 4.6 |  |  |  |  |  |
|  | **5** | 1 | 0.3 | 0.3 |  |  |  |  |  |
|  | **Total** | 259 | 100.0 |  |  |  |  |  |  |
| **3** | **1** | 88 | 41.1 | 4.9 |  |  |  |  |  |
|  | **2** | 14 | 4.3 | 1.3 |  |  |  |  |  |
|  | **3** | 38 | 25.2 | 5.3 |  |  |  |  |  |
|  | **4** | 60 | 26.2 | 4.5 |  |  |  |  |  |
|  | **5** | 5 | 3.3 | 1.8 |  |  |  |  |  |
|  | **Total** | 205 | 100.0 |  |  |  |  |  |  |
| **4** | **1** | 174 | 39.4 | 3.4 |  |  |  |  |  |
|  | **2** | 55 | 18.8 | 3.7 |  |  |  |  |  |
|  | **3** | 116 | 21.5 | 3.4 |  |  |  |  |  |
|  | **4** | 108 | 19.3 | 2.5 |  |  |  |  |  |
|  | **5** | 5 | 1.0 | 0.6 |  |  |  |  |  |
|  | **Total** | 458 | 100.0 |  |  |  |  |  |  |

**Table SB 11.** Weighted support of catchment size monitoring with history of SARS⍰CoV⍰2 infection (N-IgG) by area for probability samples (N = 981), Louisville/Jefferson County.

| Area | Support Measuring<br>Largest Areas<br>(>50,000<br>households) |  | Support Measuring<br>Smaller Sections<br>(>30,000<br>households) |  | Support Measuring<br>Neighborhoods<br>(>5,000 households) |  | Neither Support nor<br>Oppose |  | Oppose Measuring<br>Any Size |  |
| --- | --- | --- | --- | --- | --- | --- | --- | --- | --- | --- |
|  | Positive<br>%<br>(N-IgG) | Negative<br>%<br>(N-IgG) | Positive<br>%<br>(N-IgG) | Negative<br>%<br>(N-IgG) | Positive<br>%<br>(N-IgG) | Negative<br>%<br>(N-IgG) | Positive<br>%<br>(N-IgG) | Negative<br>%<br>(N-IgG) | Positive<br>%<br>(N-IgG) | Negative<br>%<br>(N-IgG) |
| 1 | 59.0 | 50.4 | - | 9.6 | 20.1 | 33.4 | 8.3 | 6.6 | 12.7 | - |
| 2 | 21.9 | 50.5 | 23.9 | 5.9 | 28.7 | 23.8 | 25.5 | 19.4 | - | - |
| 3 | 29.8 | 44.7 | - | 4.4 | 58.2 | 16.1 | 6.2 | 32.2 | 5.9 | 2.6 |
| 4 | 26.4 | 42.3 | 27.5 | 17.0 | 28.6 | 19.8 | 17.5 | 19.6 | - | 1.3 |

## Supplement C

### Wastewater Monitoring in the News, Louisville, KY

1. WBRD, Louisville health officials concerned about possible spread of COVID-19 Delta variant - Jul 7, 2021 (https://www.wdrb.com/news/louisville-health-officials-concerned-about-possible-spread-of-covid-19-delta-variant/article_27a1149e-de6d-11eb-8362-0b7fe26b39e6.html)
2. WLKY, Louisville could become only city in U.S. to document herd immunity -- with help of wastewater - Apr 15, 2021 (https://www.wlky.com/article/louisville-could-become-only-city-in-us-to-document-herd-immunity-with-help-of-wastewater/36135716#)
3. Scripps Media - University of Louisville documenting herd immunity using wastewater - May 21, 2021 (https://www.thedenverchannel.com/news/national-politics/the-race/university-of-louisville-documenting-herd-immunity-using-wastewater)
4. WAVE3, Brazilian variant of COVID-19 found in Louisville’s wastewater - May 13, 2021 (https://www.wave3.com/2021/05/13/brazilian-variant-covid-found-louisville-wastewater/)
5. UofL News, UofL receives $8.6 million from the CDC for COVID-19 wastewater research - April 14, 2021 (https://www.uoflnews.com/section/science-and-tech/uofl-receives-8-6-million-for-covid-19-wastewater-research/)
6. WDRB, California COVID-19 variant detected in Louisville - Mar 2, 2021 (https://www.wdrb.com/news/california-covid-19-variant-detected-in-louisville-as-city-preps-for-johnson-johnson-vaccines/article_a76e7b22-7b70-11eb-8992-a310c3dfa719.html)
7. Kentucky Waterways Alliance, Wastewater and surface water webinar - Jan 19, 2021 (https://zoom.us/rec/share/P84J7YFiHXfgVp6f_sDe-ainT_OmB7n2KAVc4dGip5xeDfQXVDS5pu5h70lMH4Rh.p7tOR7kjlrhWkxAM?fbclid=IwAR2QhC3V46RXEnhrPKd1mBUO4OgblRXCJUkZjC0dJAxpxpg_4Ta2X_V_bRM)
8. NY Times, Watching what we flush could help keep a pandemic under control - Nov 24, 2020 (https://www.nytimes.com/2020/11/24/magazine/coronavirus-sewage.html)
9. UofL News, MSD and UofL testing Louisville wastewater to track COVID-19 - June 18, 2020 (https://www.uoflnews.com/section/science-and-tech/msd-and-uofl-testing-louisville-wastewater-to-track-covid-19/)

